# Risk assessment of banknotes as a fomite of SARS-CoV-2 in cash payment transactions

**DOI:** 10.1101/2021.12.03.21267258

**Authors:** Jack Schijven, Mark Wind, Daniel Todt, John Howes, Barbora Tamele, Eike Steinmann

## Abstract

**Background:** The COVID 19 pandemic has triggered concerns and assumptions globally about transmission of the SARS-CoV-2 virus via cash transactions.

**Objectives:** Assess the risk of contracting COVID-19 through exposure to SARS-CoV-2 via cash acting as a fomite in payment transactions.

**Methods:** A quantitative microbial risk assessment was conducted for a worst-case scenario assuming an infectious person at the onset of symptoms, when virion concentrations in coughed droplets are at their highest. This person then contaminates a banknote by coughing on it and immediately hands it over to another person, who might then be infected by transferring the virions with a finger from the contaminated banknote to a facial mucous membrane. The scenario considered transfer efficiency of virions on the banknote to fingertips when droplets were still wet and after having dried up and subsequently being touched by finger printing or rubbing the object.

**Results:** Accounting for the likelihood of the worst-case scenario to occur by considering 1) a local prevalence of 100 COVID-19 cases/100,000 persons, 2) a maximum of about 1/5^th^ of infected persons transmit high virus loads and 3) the numbers of cash transactions/person/day, the risk of contracting COVID-19 via person-to-person cash transactions was estimated to be much lower than once per 39,000 days (107 years) for a single person. In the general populace, there will be a maximum of 2.6 expected cases/100,000 persons/day. The risk for a cashier at an average point of sale was estimated to be much less than once per 430 working days (21 months).

**Discussion:** The worst-case scenario is a rare event, therefore, for a single person, the risk of contracting COVID-19 via person-to-person cash transactions is very low. At a point of sale, the risk to the cashier proportionally increases but it is still low.

## 1. Introduction

The current SARS-CoV-2 inflicted COVID 19 pandemic has triggered concerns globally based on general assumptions about transmission of the virus via the exchange of banknotes and coins (Auer et al., 2020; Central Banking, 2020; Gardner, 2020; Roh, 2020). This was linked to general fears that any frequently touched objects such as banknotes and coins have long been suspected to serve as a transmission vehicle (fomites) of various pathogenic bacteria, parasites, fungi and viruses and now including SARS-CoV-2 (Angelakis et al., 2014; Galbadage et al., 2020; Pal and Bhadada, 2020). In the absence of evidence to the contrary, and based on the relative importance of the four normal viral transmission routes (direct transmission by droplets and touch, indirect transmission via aerosols and fomites), a diligent precautionary approach was adopted and promoted by governments and public health authorities (WHO, 2019; CDC, 2019). On top of that, some central banks also issued statements that they were temporarily quarantining or disinfecting deposited banknotes before reissuing them, or even incinerating them (Choi, 2020; Schroeder and Irrera, 2020; Yeung, 2020). These types of statements undoubtedly had an impact on the means of payments citizens decided to use for their transactions. Indeed, survey data collected by the European Central Bank in July 2020 among the general public showed that around 40% of the population of the euro area was to a certain degree concerned about the risk of being infected by handling cash. At the same time, it was seen that one of the main reasons for respondents changing their payment behaviour was simply convenience, it was also partially due to the general concern of being infected via the banknotes themselves, or via hand contact or close proximity to a cashier. It probably was also because of various governmental recommendations to pay cashless (ECB, 2020), which subsequently led to an increase in the use of other means of payment. Still, cash plays a very important role in society worldwide. As stated by the ECB (2020), it is in fact indispensable for a number of reasons, among which the social inclusion of a substantial number of citizens with limited or no access to alternative forms of payment. It is therefore in the interests of society to properly assess the risk due to exposure to SARS-CoV-2 particles via cash as a fomite in payment transactions, and not make uninformed decisions.

The US Centres for Disease Control and Prevention (CDC) has recently published a “Science brief” on the possible transmission of SARS-CoV-2 from surfaces (CDC, 2021). Here, it is pointed out that the risk of fomite-mediated transmission is dependent on the infection prevalence rate in the community, the amount of virus that infected people expel, the deposition of expelled virus particles onto surfaces (fomites), the interaction with environmental factors, the time between surface contamination and a person touching the surface, the transfer efficiency of virus particles from fomite surfaces to hands and from hands to mucous membranes on the face (nose, mouth, eyes) and, lastly, the dose of virus needed to cause infection through the mucous membrane route. In this science brief, it is concluded that the risk of fomite transmission of SARS-CoV-2 is considered low compared with direct contact, droplet transmission, or airborne transmission, whilst referring to Kampf et al. (2020) and Meyerowitz et al. (2020). Also in this science brief, references to reports are given that indicate that SARS-CoV-2 is transmitted between people by touching surfaces that an ill person has recently coughed or sneezed upon, and then immediately touching their mouth, nose, or eyes (Harvey et al., 2020, Pitol and Julian, 2020, Wilson et al., 2021). The frequency of contact with fomites as part of the risk assessment was not however mentioned in this science brief.

Recently, Todt et al. (2021) examined the stability of SARS-COV-2 on different means of payment and developed a touch transfer method to examine transfer efficiency from contaminated surfaces to fingertips. The transfer efficiencies as well as the persistence of the virus from Todt el al. (2021) indicated that the transmission of SARS-CoV-2 via contaminated coins and banknotes requires high viral loads and a timely order of specific events.

To understand this better, a quantitative microbial risk assessment (QMRA) was conducted in order to quantify the risks related to a simple but worst-case scenario where an infectious person (provider) contaminated a banknote by coughing on it, and then immediately handed it over to a second person (acceptor). The acceptor was then possibly infected by transferring SARS-CoV-2 virions with their fingers from the contaminated banknote to a facial mucous membrane. The transfer data of Todt et al. (2021) was analysed to pave the way to assess the risk of SARS-CoV-2 transmission by banknotes and coins, between two people. The risk assessment was focussed on banknotes and not on coins, because it can be assumed that coins generate a lower risk compared to banknotes simply due to their much-reduced exposed surface areas.

## 2. Methods

### 2.1 Scenario

The risk assessment comprised the following sequence of events thus defining a worst-case scenario as schematically depicted in Figure 1:

**Figure 1.**
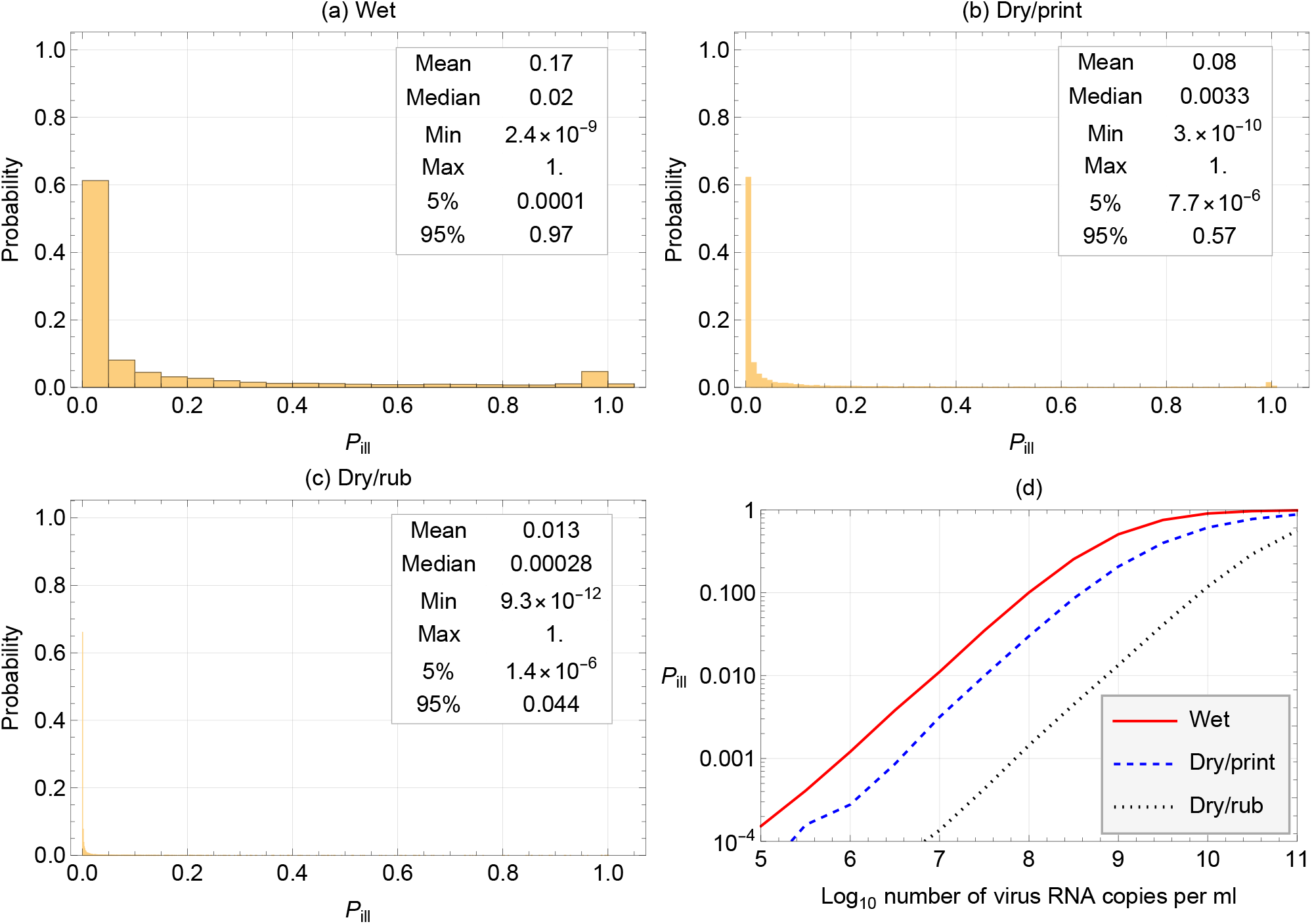
Schematic view of the contamination scenario.

1. A highly infectious person holds a banknote to make a payment and coughs upon it. The cough generates a plume of respiratory droplets and droplet nuclei containing SARS-CoV-2 virions. Part of the plume contaminates the banknote with virions.
2. The infectious person then passes the banknote to another person (acceptor). Virions are transferred from the banknote to the fingers of the acceptor via touching the freshly contaminated banknote.
3. Virions are then transferred from the acceptor’s fingers to their eyes, nose or mouth by touching. This number of transferred virions is considered to be the dose.

Note that the assessment did not cover the many other likely permutations of the possible transfer route, i.e., the possible intermediary role of wallets, cash machines, pockets, other precautionary measures such as face masks, hand cleaning and the use of rubber gloves as these routes most probably would lead to a lower amount of virions transferred.

The risk characterisation entailed the dose response relationship, expressing the risk per event as described in the exposure assessment, and a sensitivity analysis.

Because only short time frames were considered in this sequence of events, and SARS-CoV-2 has been observed to remain infectious in aerosols and on banknotes for hours (Fears et al. 2020; van Doremalen et al. 2020, Todt et al. 2021), decay over time was not modelled in this study. Due to the drying of virus-containing droplets on a surface, virus inactivation proceeds initially faster as shown by Todt et al. (2021). But in a timeframe of seconds to a minute this does not play a role in this assessment and has not been taken into account. The lower transfer efficiencies under dry conditions may be due to picking up virions less efficiently in combination with the numbers of virions that were already reduced due to the drying.

This assessment concludes with an estimate of the likelihood of this depicted worst-case scenario while putting the risk assessment in perspective for the general risk per person per day, for both person-to-person and points-of-sale cash transactions.

### 2.2 Risk assessment

#### Virus concentration

It was assumed that the viral concentration *C* [virus RNA copies/mL] in droplets from a cough were the same as determined in throat and nasal swabs from infected persons. To this end, viral concentration data collected during the pandemic before July 2020 as published by Schijven et al. (2021) were used. Here, only the onset of symptoms when the concentrations are highest was considered. Table 1 lists the corresponding parameters.

**Table 1.**
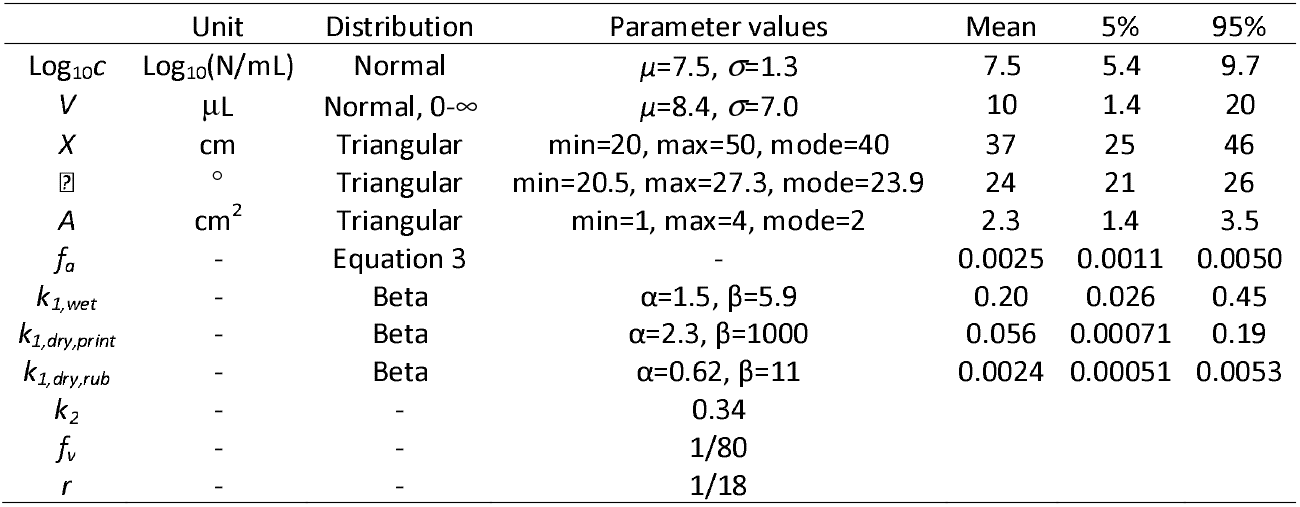
Parameter values.

|  | Unit | Distribution | Parameter values | Mean | 5% | 95% |
| --- | --- | --- | --- | --- | --- | --- |
| $\text{Log}_{10}C$ | $\text{Log}_{10}(\text{N/mL})$ | Normal | $\mu=7.5, \sigma=1.3$ | 7.5 | 5.4 | 9.7 |
| $V$ | $\mu\text{L}$ | Normal, $0-\infty$ | $\mu=8.4, \sigma=7.0$ | 10 | 1.4 | 20 |
| $X$ | cm | Triangular | min=20, max=50, mode=40 | 37 | 25 | 46 |
| ∅ | ° | Triangular | min=20.5, max=27.3, mode=23.9 | 24 | 21 | 26 |
| $A$ | $\text{cm}^2$ | Triangular | min=1, max=4, mode=2 | 2.3 | 1.4 | 3.5 |
| $f_a$ | - | Equation 3 | - | 0.0025 | 0.0011 | 0.0050 |
| $k_{1,wet}$ | - | Beta | $\alpha=1.5, \beta=5.9$ | 0.20 | 0.026 | 0.45 |
| $k_{1,dry,print}$ | - | Beta | $\alpha=2.3, \beta=1000$ | 0.056 | 0.00071 | 0.19 |
| $k_{1,dry,rub}$ | - | Beta | $\alpha=0.62, \beta=11$ | 0.0024 | 0.00051 | 0.0053 |
| $k_2$ | - | - | 0.34 | | | |
| $f_v$ | - | - | 1/80 | | | |
| $r$ | - | - | 1/18 | | | |

#### Total volume of droplets in a cough

Data on numbers and size distributions of expiratory droplets expelled during coughing were kindly provided by Chao et al. (2009), who measured droplet size using the interferometric Mie imaging. In that study, measurements were conducted at 1 cm in front of the mouth. Therefore, evaporation of the droplets (which would decrease their volume and, hence, increase the concentration of virions in the droplets) was not considered. Because of the 20 cm – 50 cm distance between the mouth of the infectious person and his/her hand holding a banknote, the whole spectrum of expelled droplets as found by Chao et al. (2009) was considered. Chao et al. (2009) used a factor of 18.94 to obtain a mean total droplet count of 2085. Chao’s data encompassed variability in measurements from ten healthy volunteers. For each volunteer *i*, data was collected in seventeen size bins of different diameters ranging from 1 to 2000 µm, that were converted to mean volumes *V*_*i,j*_ [millilitres] per size bin *j*:

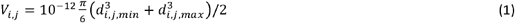

where *d* is the diameter [µm] and *min* and *max* designate minimum and maximum values per size bin, respectively.

The total volume of droplets from a cough per volunteer, *V*_*i*_ [ml] was calculated by summing *n*_*i,j*_ samples of *V*_*i,j*_ for each V_i_:

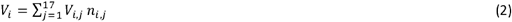

Finally, a normal distribution that was truncated between zero and infinity was fitted to the ten *V*_*i*_ values.

#### Caught fraction of projected cough plume

It was assumed that at the moment of coughing, the infectious person held a banknote in their hand at a distance X from the face, and only a fraction of the cough plume directed to the banknote actually contaminated the banknote. Subsequently, the exposed person accepted the banknote and touched part of its surface area with their fingertips where transfer of virions from the banknote to the fingertips could take place. The fraction of the projected cough plume on the banknote that is touched by the fingertips and thereby caught by the exposed person, *f*_*a*_, was calculated as:

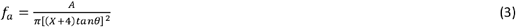

where *A* is the surface area [cm^2^] of the fingertips touching the banknote, *X* is the distance [cm] between the mouth of the infectious person and the banknote at the time of coughing. Distance *X* was augmented by 4 cm to include the distance from the throat (the origin of the plume) to the mouth opening. ⍰ is the opening angle of the cough plume (Bourouiba et al.,2014). *A, X* and ⍰ were all modelled as triangular distributions. See Table 1 for the corresponding distribution parameter values.

#### Transfer efficiencies from banknote to hand

The transfer efficiency data of infectious SARS-CoV-2 virions from surfaces of banknotes and coins to artificial skin as reported by Todt et al. (2021) were analysed by means of a multivariate linear regression for the relationship between the transfer efficiencies and the input concentrations, types of surfaces, and transfer methods and conditions. (Input concentrations and transfer efficiencies: log base 10; types of surfaces: €0,10 coin, €10 banknote, PVC, steel; transfer condition: wet = immediate contact after inoculation, dry = after desiccation of the droplet; transfer method: by finger print of finger rubbing; regression analyses: R (version 3.5.2 (2018-12-20) - “Eggshell Igloo”) and lm (Chambers, 1992; Wilkinson and Rogers, 1973)). Note that data for input concentrations less than 2320 infectious virions per ml were excluded from the analysis, because those values as well as the output concentrations (after transfer) were near or below the detection limit and did not allow to determine a transfer efficiency. The model with the lowest Akaike’s Information Criterion (AIC) was selected using the step-function (parameter k=3.84). For graphical presentation of the data package ggplot2 was used (Wickham, 2016). Transfer efficiencies were grouped along the significance of the effects of type of surface, transfer condition and method. For each group of transfer efficiencies, a Beta distribution was fitted and labelled *k*_*1,group*_. Note that it was assumed that the transfer efficiencies from Todt et al. (2021) for infectious virions also applied to virus RNA copies.

The rate of hand contact with target facial membranes (mouth, eyes, nostrils) was estimated to be on average 15.7 per hour (Nicas and Best, 2008), i.e., once every 4 minutes, and, therefore, frequent enough to justify almost immediate transfer to the facial membranes after having touched the contaminated banknote. For transfer efficiency, *k*_*2*_, of SARS-CoV-2 from contaminated fingers to the face, *i*.*e*., lips, nose, eyes, the value 0.34 for transfer of bacteriophage PRD1 was taken from Rusin et al. (2002). Bacteriophage PRD1 is an environmentally persistent virus, which is strongly negatively charged, and, therefore, can be considered as a conservative model virus regarding virus persistence and attachment to surfaces (Schijven and Hassanizadeh, 2000).

#### Dose and risk

The dose *D* was computed as follows:

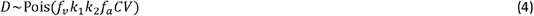

Where *f*_*v*_ =1/80 and is a conservative estimate of the fraction of intact virions that are able to infect specific cells in a tissue culture (Schijven et al., 2021).

Finally, the risk of contracting COVID-19, *P*_*ill*_, was computed using the exponential dose response model:

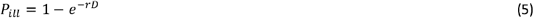

where *r* is the preliminary dose response parameter proposed by Haas (2021). Haas (2021) recommended to use the dose response data for human coronavirus 229E as representative for SARS-CoV-2. For human coronavirus 229E, it was found that each plaque forming unit (pfu) has a probability of r=1/18 of causing illness symptoms.

#### Fitting of distributions and Monte Carlo simulation

Fitting distributions to data of *V* and *k*_*1,group*_ and drawing 10 000 Monte Carlo samples of *C, V, A, X*, ⍰ and *k*_*1,group*_ were conducted in Mathematica (version 12.3.0.0, Wolfram Inc, Champaign, IL, USA).

## 3. Results

Table 3 lists the numbers of virus RNA copies present in a single cough (“coughed”), caught on a banknote area of a size covered by fingertips (“caught on fingertip area”), then actually transferred to the fingertips and, finally to a mucosal site of the facial area (“dose”). These numbers varied by many orders of magnitude, which stemmed from a combination of the large variation in virus concentration in patient material and the large variation in volume of coughed droplets. The numbers caught on the banknote on a fingertip-sized area were about 400 times lower than the numbers present in a single cough. Under wet conditions, the numbers transferred to the fingertips were lowered by about 5 times. Under dry conditions, the virus RNA copy numbers were reduced 20 and 400 times by printing and rubbing, respectively. The amount transferred from the fingertip to the facial area, the dose, was about one-third of the numbers on the fingertips. At the end, the dose was on average about 5,000, 25,000 and 500,000 times lower than the numbers of coughed virions for the wet, dry/print and dry/rub scenarios, respectively.

Figure 2 presents the boxplots of the transfer efficiency data according to the type of surface and transfer under wet and dry conditions by printing and rubbing, and shows that the transfer efficiencies under wet conditions were higher than under dry conditions. It can also be seen that under wet conditions, transfer efficiencies were similar for printing and rubbing, but that under dry conditions, transfer efficiencies were lower for rubbing than for printing. According to the statistical analyses of the transfer efficiency data, input concentration and the type of surface did not significantly affect transfer efficiency, but, dry/wet conditions and printing/rubbing did significantly affect transfer efficiency. Under wet conditions, transfer efficiencies were on average 10 times higher than under dry conditions and transfer efficiencies were on average 3 times lower for rubbing compared to printing (See estimated coefficients in Table 2). Therefore, for the risk assessment, the beta distributions were fitted to the transfer efficiencies *k*_*1*_ of the €10 banknote under wet conditions, regardless of printing or rubbing and for the dry conditions accounting for printing and rubbing. The corresponding parameters of the three *k*_*1*_’s are listed in Table 1.

**Table 2.**
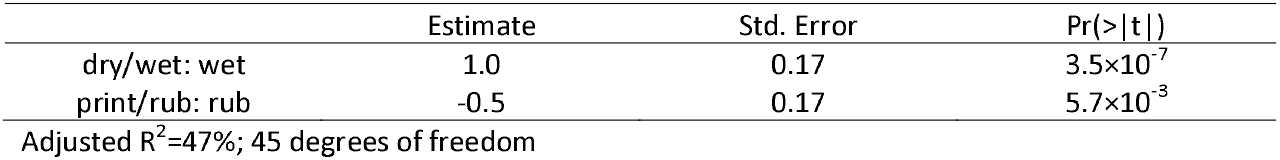
Parameter estimates from regression analysis of the log^10^ of the transfer efficiencies Adjusted R^2^=47% 45 degrees of freedom.

**Table 3.** Numbers of virus RNA copies in the coughed plume, caught on fingertip area, transferred to fingertips and dose according wet, dry/print and dry/rub scenarios.

|  | Mean | Min | Max | Median | 5% | 95% |
| --- | --- | --- | --- | --- | --- | --- |
| Coughed | $2.5 \times 10^7$ | 2 | $2.0 \times 10^{10}$ | $2.5 \times 10^5$ | $1.6 \times 10^3$ | $4.3 \times 10^7$ |
| Caught on fingertip area | $6.2 \times 10^4$ | 0 | $4.3 \times 10^7$ | $5.6 \times 10^2$ | 3 | $1.0 \times 10^5$ |
| Wet: transfer to fingertips | $1.4 \times 10^4$ | 0 | $1.5 \times 10^7$ | $8.4 \times 10^1$ | 0 | $1.7 \times 10^4$ |
| Wet: dose | $4.7 \times 10^3$ | 0 | $5.1 \times 10^6$ | $2.9 \times 10^1$ | 0 | $5.8 \times 10^3$ |
| Dry/print: transfer to fingertips | $3.1 \times 10^3$ | 0 | $5.6 \times 10^6$ | $1.3 \times 10^1$ | 0 | $3.8 \times 10^3$ |
| Dry/print: dose | $1.0 \times 10^3$ | 0 | $1.9 \times 10^6$ | 5 | 0 | $1.3 \times 10^3$ |
| Dry/rub: transfer to fingertips | $1.5 \times 10^2$ | 0 | $1.4 \times 10^5$ | 1 | 0 | $2.0 \times 10^2$ |
| Dry/rub: dose | $5.0 \times 10^1$ | 0 | $4.8 \times 10^4$ | 0 | 0 | $6.9 \times 10^1$ |

**Figure 2.**
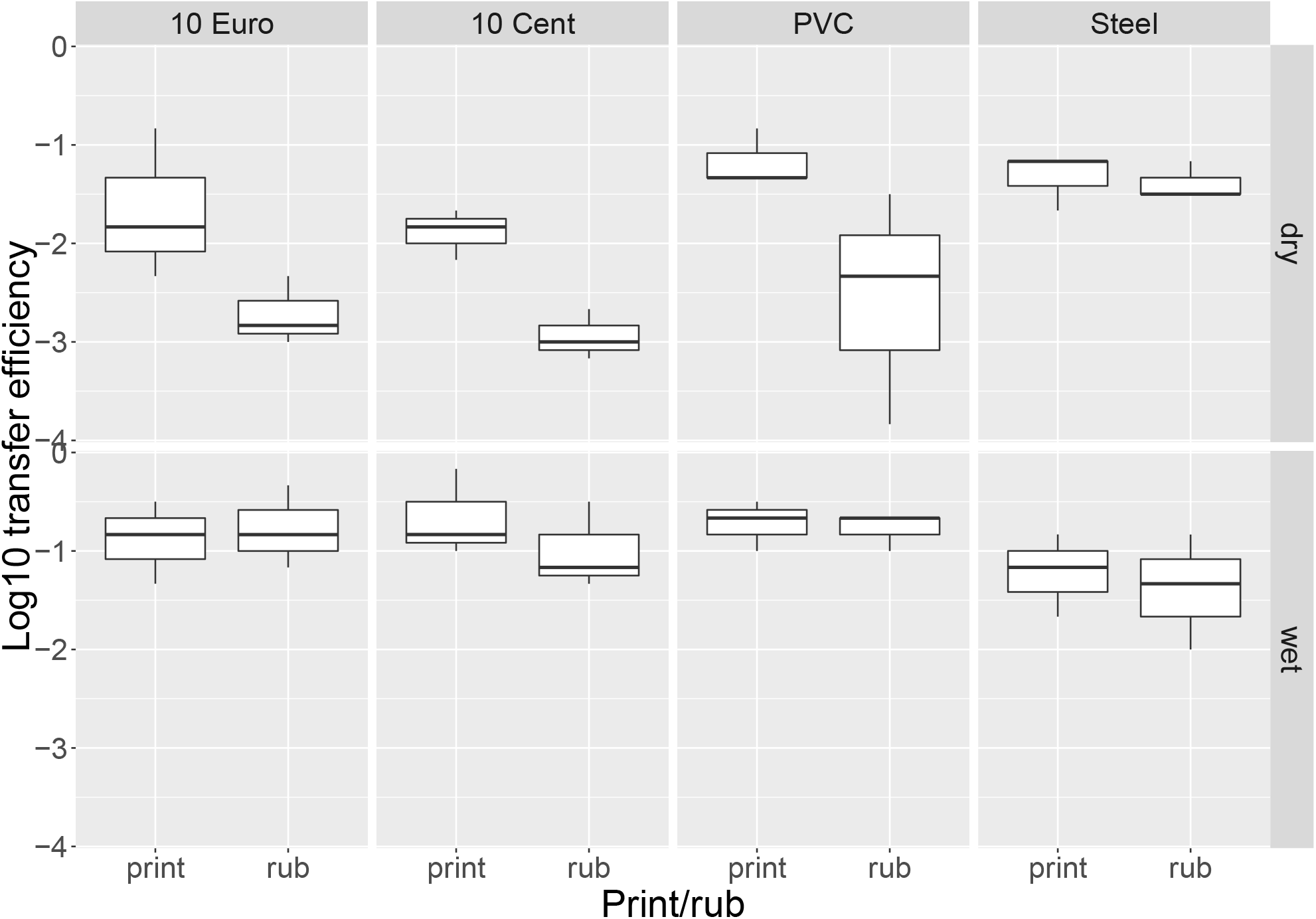
Boxplots of the log^10^ transfer efficiency data according to the type of surface and transfer under wet and dry conditions by printing and rubbing. Data from Todt et al. (2021).

Figures 3a – 3c depict the corresponding mean risks of 0.17 (=1/6), 0.082 (=1/12) and 0.014 (=1/71) per cash transaction under the worst-case scenario, which is assuming an infectious person immediately handed over a heavily contaminated banknote followed by wet, dry/print and dry/rub transfer conditions, respectively. The risk distributions spanned the range from zero to one due to the wide ranges of the transferred numbers of virus particles. These risk estimates can also be expressed as contracting on average COVID-19 once per 6, 12 and 71 cash transactions carried out under the worst-case scenario for the wet, the dry/print and the dry/rub transfer conditions, respectively.

**Figure 3.**
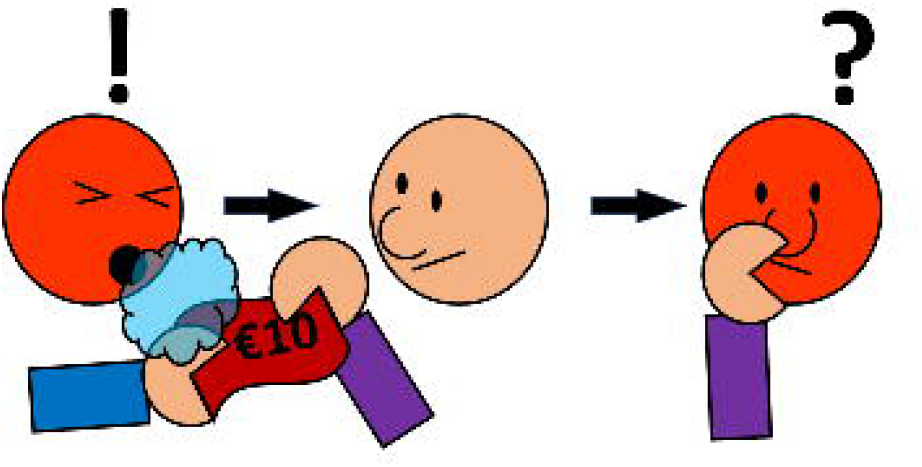
**Histograms (a, b, c) of the distribution of the risk *P*_*ill*_ of contracting COVID-19 according to three transfer scenarios (wet, dry/print and dry/rub transfer) and (d) the mean risk *P*_*ill*_ of contracting COVID-19 as a function of the number of virus RNA copies per ml mucus for each the three transfer conditions in the worst-case scenario.** **The probability that an infectious person at the onset of symptoms is expelling at least 10^5^, 10^6^, 10^7^, 10^8^, 10^9^, 10 or 10^10^ RNA copies/mL is 100%, 88%, 66%, 36%, 13%, 2.7% and 0.34%, respectively, based on virus load data in the beginning of the pandemic.**

Figure 3d depicts the mean risk *P*_*ill*_ of contracting COVID-19, as a function of a given number of virus RNA copies per ml mucus for each of the three transfer scenarios over the concentration range from 10^7^ to 10^11^ virus RNA copies per ml. In the legend, the corresponding percentiles are given for having at least a specific virus concentration. From these curves, it can be seen that the risk level of 1% will be exceeded at a virus concentration of about 10^7^, 6×10^7^ and 10^9^ virus RNA copies per ml for the wet, dry/print and dry/rub transfer scenarios, respectively. The risk level of 10% will be exceeded at a virus concentration of about 10^8^, 6×10^8^ and 10^10^ virus RNA copies per ml for the wet, dry/print and dry/rub transfer scenarios, respectively. In summary, under the worst-case scenario, in which an infectious person immediately handed over a contaminated banknote, these curves imply that in the wet transfer scenario, with an about average virus concentration of 10^8^ virus RNA copies per ml, the risk can be considered high, which is no surprise given the worst-case conditions. But in the dry/rub scenario, a high virus concentration as produced by a super spreader expelling virus at a concentration of 10^10^ virus RNA copies per ml would be required to achieve the same risk level.

## 4 Discussion

The current study aimed to assess, using a number of worst-case assumptions, the risk of becoming ill through using cash in a transaction with another person, which acts as a fomite of SARS-CoV2 during cash payment transactions. The worst-case assumptions are as follows:

1. The person that expelled the SARS-CoV-2 virions was at the onset of symptoms, when virion concentrations in the expelled droplets are at their highest.
2. The cough was aimed such that the cough plume completely encompassed the banknote which was held normal to the direction of the cough.
3. Contact between the banknote and fingertips and subsequently facial area was immediate, implying inactivation of virions was negligible.
4. 1 PFU corresponded to a conservative estimate of one per 80 virus RNA copies (Schijven et al., 2021). The latter in combination with the prudent preliminary dose response parameter of 1/18 (Haas et al., 2021), corresponded to 1/1440 virus RNA copies on average leading to illness, which was not too conservative after all because it matched the number of viral genomes needed to initiate infection of about 1000 (1-5000) as reported by Popa et al. (2020) based on a transmission network with thirty-nine transmission events. Basu (2021) estimated an infectious dose of about 100 virions to explain a superspreading event.
5. Current preventative measures such as hand cleaning and face masks and physical barriers between cashiers and the public were not taken into account.

To determine the volume of droplets in a single cough using the data from Chao et al. (2009) variability in numbers of coughed droplets between persons was accounted for. Jones (2020) also used the Chao data but accounting for variability by using Poisson variance for each diameter class, which is a much narrower distribution. In contrast, Duguid (1945) reported much wider ranges of expelled particles of 1 – 100 micrometres by a lip cough of 490 – 16.000 or a tongue-teeth cough of 1500 – 52000.

According to the statistical analysis of the transfer efficiency data, the type of surface was apparently an insignificant factor, nevertheless, risk estimates for a scenario in which a €1 or €2 coin was handed over should be much lower simply because the fingertips of the payer shield most of the coin surface area against virus contamination from a cough.

The risk assessment demonstrated that risks of contracting COVID-19 could be high in the worst-case events as described. Transfer efficiencies were derived from the experimental data from Todt et al. (2021), in which droplet spots of 10 ul (diameter of about 5 mm) were deposited on the different surfaces. Desiccation has been shown to reduce the numbers of virions by two orders of magnitude after 7 hours of time (Todt et al., 2021). In the contamination event, the amount of mucus caught on the banknote is likely dispersed over much smaller droplets, and, hence, desiccation may happen much faster.

From equation 4, it is obvious that dose *D* is directly proportional to the concentration of virus RNA copies *C*, the volume of droplets in a single cough *V*, the fraction of the cough caught by the area covered by fingertips *f*_*a*_, the transfer efficiencies *k*_*1*_ and *k*_*2*_, and, finally, the fraction of infectious viruses *f*_*v*_. Table 1 shows that the 5-percentile 95-percentile range of C spanned four orders of magnitude, but more or less one order for all the other parameters. Therefore, the variability in *D* is by far determined by *C*. Figure 3d depicts the mean risk *P*_*ill*_ of contracting COVID-19 as a function of the number of virus RNA copies per ml mucus for each the three transfer scenarios over the concentration range from 10^7^ to 10^11^ virus RNA copies per ml. Considering 10^10^ virus RNA copies per ml representing a super spreader event (Schijven et al., 2021), mean values of *P*_*ill*_ become high for all three transfer scenarios. The viral load of the Delta variant might be at this level because there are indications that the viral load of the Delta variant is 1000 times higher than the initial variants of 2020 (Li et al., 2021).

The virus transmission route that was chosen, i.e., coughing onto a banknote and then immediately passing it to another person by hand, can be assumed to happen only very rarely in public life because it is neither socially acceptable nor courteous, and people would normally avoid such an action. During the pandemic, it probably became even less likely due to hygienic measures, social distancing, wearing of masks and increased awareness. Evaluation of the likelihood of the depicted event entails analysing the frequency of cash transactions, prevalence of SARS-CoV-2 infection as well as physical/biological susceptibility to SARS-CoV-2 infection. According to a study on the payment attitudes of consumers in the euro area in 2019, 1.2×10^11^ cash transactions (1.1×10^11^ point- of-sale and 8.0×10^9^ person-to-person) were made involving a value of €2.0×10^12^ (ECB, 2020). This amounts to an average of €17 per cash transactions and may be interpreted that at least one banknote is involved in each cash transactions. The same study estimated that on average in the euro area, consumers made 1.1 cash transactions and 0.4 card transactions per person per day (ECB, 2020).

As previously stated, in the depicted worst-case event, the risk of contracting COVID-19 under wet transfer conditions from a banknote was estimated to be 1 per 6 events. In reality, not every infectious person is at the onset of symptoms when emission of virions is at its highest. About 10 to 20% of people with COVID-19 cause 80% of subsequent infections – which may lead to so-called superspreading events – while 60-75% of people with COVID-19 infect no one else (Bi et al., 2020; Chen et al., 2021; Endo et al., 2020; Goyal et al., 2021). Most people with COVID-19 infect no one because they expel little – if any – infectious SARS-CoV-2 when they talk, breathe, sing or cough, but highly infectious individuals on the other hand have high concentrations of the virus in their airways, particularly the first few days after developing symptoms, and can expel tens to thousands of infectious virus particles per minute (Chen et al., 2021). Interpreting this heterogeneity in transmissibility and shedding of SARS-CoV-2, in which at maximum about one fifth of infected persons transmit high loads of virus particles, it may be assumed that estimated risks are at least five times lower than those estimated for the worst-case scenario with a person at the onset of having symptoms. Assuming a local prevalence of SARS-CoV-2 infections of 100 per 100,000 persons, which falls well within an area classified as orange in the standard publicised ECDC charts (ECDC, 2021), together with the knowledge that there are 1.1 cash transactions per person per day, this would imply that a person-to-person cash transaction with an infectious person would occur with a probability of 1.1/1000 per day. For a single person, this leads to a risk estimate of contamination of once per 5×1000/(0.17×1.1×0.7) = 39,000 days (107 years) for person-to-person cash transactions.

Also, hardly any person will cough on a banknote when paying, therefore, this risk estimate must actually be much lower than once per 39,000 days for the average citizen. How much lower because of this is unknown. Additionally, when taking into account that currently in European countries a substantial part of the population has now acquired immunity by infection or vaccination, this risk can now be proportionally lowered even further as time progresses. A second conclusion is that in the general populace, assuming 100 infectious persons per 100,000, there will be much less than 100,000×1/5×0.17×1.1×0.7/1000 = 2.6 expected cases per 110,000 transactions (100,000 persons) per day.

Point-of-sale and person-to-person cash transactions differ significantly in frequency and exposure levels and therefore should be considered separately. At any point of sale, a cashier at a supermarket or similar shop may be dealing with cash transactions many more times per day, and is therefore, more exposed to the risk of touching contaminated banknotes and objects. At an average point of sale, with a cashier conducting 100 cash transactions per day, the risk for the cashier with no protection is obviously proportionally increased compared to the risk per single person-to-person cash transactions to a rate much lower than once per (39,000×1.1/100=) 430 working days (21 months). This again emphasises that it is important that adequate precautions are taken at points of sale to protect the cashier. Having an infectious cashier must obviously be avoided at all times, emphasising the importance of the public messages for regular testing and that every person with symptoms must self-isolate.

To conclude, the overall risk of transmission via banknotes between members of the general public with no protection is very low (much less than once every 39,000 days). The worst-case scenario as analysed in this paper may be a high-risk event, but it is also a rare event. Obviously, cashiers are more at risk than the public (much less than once every 430 days) even if still with a low risk to contract COVID-19 through exposure to SARS-CoV-2 via cash. In any case, this should not prevent from following the national and international guidelines in such circumstances.

Additional research is necessary to assess if the risk increases significantly because of potentially newer virus variants.

## Data Availability

All data produced in the present study are available upon reasonable request to the authors

## Acknowledgements

We thank Ana Maria de Roda Husman, Alvin Bartels, Erwin Duizer, Arno Swart and other anonymous reviewers for their valuable comments on this work.

